# Immunisation Decision-Making and Barriers to Vaccine Uptake among Children Under-5 in Low-Resource Settings

**DOI:** 10.1101/2025.05.21.25328058

**Authors:** Gbadebo Collins Adeyanju

## Abstract

**Background:** Since the turn of the millennium, childhood immunisation against preventable infectious diseases in sub-Saharan Africa (SSA) has experienced unprecedented growth. However, immunisation coverage remains a fundamental challenge in the region, which still has the highest under-five mortality rate in the world, due to wide-ranging drivers that have complicated interventions. Low immunisation coverage will continue to plague the region unless the knowledge-gaps associated with childhood immunisation decision-making are identified, studied, measured and addressed. Therefore, the aim of this study is to assess the factors that influence immunisation decision-making among caregivers of children under-5 years old, and to understand the behaviours that shape these influences.

**Methods:** The study used qualitative methods such as focus group discussions. Participants were caregivers of children under-5 years old in Nigeria. Simplified cluster sampling approach was used to select caregivers in four geographical clusters. A minimum of seven caregivers from each cluster were purposively included. Data were analysed deductively using meta-aggregation approach.

**Results:** The study findings show that caregivers immunisation decision-making are mainly motivated by: inadequate knowledge about childhood immunisation, especially the conflict between vaccine-preventable and non-vaccine-preventable diseases; masculinity (attitudes of fathers or men can help or hinder immunisation); the gender of child (the perception of weaker versus stronger sex); misinformation about immunisation, especially the perception that it is family planning through the backdoor). Other influences include exploitation of caregivers by healthcare workers; incessant stock-outs of vaccines leading to complacent behaviour associated with vaccine hesitancy; ineffective communication about immunisation schedule and poor reminder systems; religious beliefs; poor attitudes of healthcare workers and more.

**Conclusion:** The factors that influence immunisation decision-making in low-resource settings and the motivations that shape these behaviours are largely psychological, sociocultural, behavioural, health system and structural. Designing interventions that address the root causes of gender inequity must start with the attitudes of men and the socio-cultural practices that enable them. Furthermore, the sandwich model for addressing vaccine misinformation can be effective in countering myths and conspiracies about vaccines.

## INTRODUCTION

Since the millennium, childhood vaccination coverage for preventable infectious diseases in Sub-Sahara Africa (SSA) have received unprecedented growth [1]. The efficacy of vaccines has saved over 50 million lives in the SSA region since the introduction into the National Programme on Immunisation (NIPs) in the 1970s [2]. For every infant life within this period, about 60 years of life are lived [2]. However, despite these remarkable public health breakthroughs, acceptance and uptake still have steep height to climb in the region, as it accounts for the highest under-five mortality globally and 40% of the total deaths within this age bracket [3]. Vaccination coverage in SSA have stagnated at pre-pandemic level, and in some cases, declined [4-6]. About one in every five children in SSA have not received basic vaccines, which results in more than 30 million children under-five years suffering from vaccine-preventable diseases (VPDs) every year in the region, and out of these, more than half a million die annually, thereby constituting about 58% of global VPD-related deaths [7]. More so, the top 10 countries with the largest contribution of zero-dose children and account for over 80% of the global zero-dose children are in the SSA; they include Nigeria (30.1%), Ethiopia (14.1%), Democratic Republic of the Congo (9.5%), Angola (7.2%), United Republic of Tanzania (3.6%), Madagascar (3.4%), Mozambique (3.6%), Mali (2.4%), Chad (2.9%), and Cameroon (3.1%) [7].

The coverage of childhood immunisation in Nigeria varied across states and regions, but generally showed the need for more efforts, because immunisation coverage in Nigeria is still below the 90% target, thereby putting a significant number of children at risk of VPDs outbreak [8.9]. Nigeria is among the countries in SSA that have third dose of the Diphtheria, Pertussis, and Tetanus (DPT3) coverage lower than 80% and the country with the highest number of children not vaccinated for DTP1 (zero-dose), i.e., over 30% of zero-dose children comes from Nigeria alone [7].

In Nigeria, basic childhood immunisation includes: one dose of Bacillus Calmette Guérin (BCG); three doses of Diphtheria-Pertussis-Tetanus (DPT); three doses of Oral Polio Vaccine (OPV), one dose of Measles among others [10]. The routine immunisation of children that is being implemented by the Expanded Program on Immunisation (EPI) is carried out using regiment such as: BCG (administered at birth or as soon as possible after birth); OPV (administered at birth and at the six, 10, and 14 weeks of age); DPT (given at the six, 10, and 14 weeks of age); Hepatitis B (given at birth, sixth and 14th weeks); measles is given at 9 months of age; yellow fever is administered at 9 months of age; and Vitamin A at 9 months and 15 months of age [11].

There are several drivers of childhood immunisation decision-making in Nigeria. For instance, age, region, education, wealth index, and the number of visits to antenatal care (ANC) clinic are some major determinants of the completion of childhood immunisation [5]. Other factors are associated with the number of doses of childhood vaccines taken including maternal age and education, as well as income status, ANC attendance, employment, use of skilled birth attendants, religion, poverty, literacy, and more [10]. Immunisation behaviour among caregivers is also influenced by social factors like spousal opposition, institutional factors like health system problems, and cognitive factors like concerns about vaccine safety [12]. Psychological antecedence such as confidence, complacency, constraints, calculation, collective responsibility, religious beliefs, rumours and more have shape behaviour of caregivers when it comes to childhood immunisation [13-16]. Other factors noted at the family and community level that enabled low demand for immunisation includes lack of understanding of its value, perception of immunisation, inadequate cold chain equipment in public healthcare facilities, political crises affecting communities, vaccines availability and fear, among others [17-20].

In the SSA, several studies have associated the low vaccine uptake phenomenon to misinformation, gender disparity, masculinity factors among others [14-16,21-25]. However, there is dearth of empirical and in-depth research on these factors and the understanding of how they drive these outcomes in households’ vaccination decision-making in low-resource settings such as Nigeria. Recent studies showed that the intention to vaccinate new-born drops when caregivers believe in rumour or misinformation, especially that vaccines can cause infertility or are designed to reduce population [16,21,20,23]. Also, male partners or father’s opinion and actions are crucial in overall household decisionmaking, especially in SSA, where some studies have cited them as being not enthusiastic about vaccinating children [3,14,21]. Their knowledge, attitude and/or belief shapes household’s decision on whether a child is vaccinated or not. The disparities in immunisation coverage between boys and girls have also shown how gender of children could influence vaccination decision-making [26-28]. This has also been true outside the SSA such as Bangladesh, where girls are less likely to be fully vaccinated than boys [28].

Even where successes have been recorded, in terms of immunisation coverage and sizeable uptake, the erosion of the earlier made progress in the last decade owing to vaccine hesitancy and other wide-ranging drivers among other unclassified determinants in the SSA and Nigeria makes intervention even harder [15,16,21,22,24,29-31]. Low vaccination demand and disproportionate infant mortality would continue to plague the region if the scientific evidence needed for designing effective interventions to address childhood vaccination decisionmaking are not empirically understudied, identified, measured and addressed.

The reasons for low immunisation among caregivers are often complex, multi-faceted, context-specific, and as mentioned earlier, generally include social, institutional, psychological and cognitive factors, some of which underscores vaccine hesitancy [12]. No doubt vaccine hesitancy is still a significant theme affecting vaccination uptake, and consequently its coverage in Nigeria [14,21,30]. Lack of trust in public institutions (16%), and the prevalence of vaccine conspiracy theories mostly related to religion and biotechnology (46%), especially in the wake of COVID-19 vaccine, are identified triggers associated with vaccine hesitancy in Nigeria [32]. Meanwhile in the case of zero-dose and under-immunized children, vaccine uptake faces several interrelated barriers like gender, poverty, geographic access, service experience, and the non-alignment of national immunisation program with needs of vulnerable people [8,9]. These barriers constitute an impasse that reinforce the tendency of caregivers not to immunize their children.

Therefore, the goal of this study is to assess the underlining influence of immunisation decision-making among caregivers of children under-5 and factors that shapes those behaviour.

## METHODS

The study used a qualitative design in form of Focus Group Discussion (FGD) method. The study was conducted according to the guidelines and approved of the Nigeria’s health research ethics committee (ref. no.: FHREC/2023/01/48/04-03-23). Written informed consent was obtained from all participants.

### Sampling design

The study sample were caregivers of children Under-5 who were recruited purposively in four predefined stratified clusters, out of which eligible participants were aggregated in form of group meetings. The primary target group were mothers/fathers or legal guardians (of children between 0 - 5 years old) here referred to as caregivers in Nigeria. The sample population was stratified into four clusters using simplified cluster sampling approach [33,34]. One community from each cluster was randomly selected from which participants were pur-posively drawn. The selected communities from where participants were drawn have adequate geographical representation and health facilities providing immunisation services for at least three years prior to the study. At least seven caregivers whose children were Under-5 were included from each of the clusters, generating a total of 28 participants.

Participants were representative for age, gender, socio-economic status, religion and region of origin in Nigeria. Each participant received an information sheet stating the goals and expected outcomes of the study. This was accompanied by an Informed Consent Form, which was signed by all study participants. The FGDs were audio recorded and explored perspectives of caregivers on behavioural influence of childhood immunisation decision-making.

### Themes explored

The FGDs were guided by a carefully designed semi-structured instrument. It explored themes such as: *knowledge about childhood immunisation* (“What comes to your mind when one talks about childhood immunisation?”); *immunisation demand* (“In your experience, how will you consider current immunisation acceptance?”); *factors driving low immunisation coverage* (“In your view, what could be factors responsible for low immunisation uptake and why?”); *Importance of vaccines to a child’s health* (“How important do you think vaccines are to a child’s health? Follow-up: How important do you think for a child to received none, some or all of these vaccines?”); *masculinity* (“When it is time for children to get vaccinated, the mother would need permission to take the child to the clinic for immunisation? Followup: How does the final decision on child’s immunisation rests on the fathers? Why is his permission important? Is it because of culture, fathers are head of the house, fathers better understand immunisation, God/Allah commanded it?”). Others are: g*ender disparity: boys versus girls* (How do you see immunisation between boys and girls…do you think immunisation is more important for girls than boys or vice versa…Why do you think so?). *Misinformation* (“What do you think about all the information going around about immunisation? Follow-up…some vaccines generally contain chemicals that are harmful…prayers are more effective in preventing diseases…some vaccinations are designed to reduce our population). *Religious beliefs* (“How would you consider religious beliefs and immunisation…does it go well with religious beliefs?”). *Immunisation schedule* (“Nigeria has a schedule of vaccines for children. Do you think people want their children to receive none, some or all childhood immunisation and why?”). *Immunisation intention* (“Has anyone here received Hepatitis B vaccine? If so, why did you take it? If not, would you like to receive vaccination for Hepatitis B?”). *Impact of COVID-19 on immunisation services* (“Has the COVID-19 pandemic affected your views about immunisation generally. If so, why? If not, why?”).

### Data analysis

The data were analysed using meta-aggregation approach, which summarizes data in a stepwise process and developed themes based on predefined concepts [35,36]. After analysing each individual transcript (first-order data), confluence and deviate views based on insights sorted were identified (second-order data) and categorised based on the aggregation of subject outcomes (third-order data). Similar process sufficed for undefined themes that emerged inductively from the data as well. All discussions were audio recorded and subsequently transcribed verbatim. The transcribed data were coded based on given participant’s identifiers (RS1 – RS7) and corresponding to the clusters (f1 – f4) where they were obtained e.g., f1001-7, f2001-7, f3001-7 and f4001-7.

## RESULTS

### Drivers of Low Immunisation Uptake

### Inadequate knowledge about childhood immunisation

Caregiver’s knowledge of immunisation services was mixed or at best inadequate. There is a conflicting understanding of what is and is not vaccine-preventable, with most caregivers believing that all childhood diseases can be prevented by immunisation.

> “No parent wants to see their child deformed. If you have got a polio-ridden child, while other children are strong and healthy, you will not be happy” f2RS1; “I think immunisation is an act of making someone resistant to infectious diseases”…f1RS1. “I will say it is a kind of medicine given to a child to prevent any kind of disease”…f2RS1. “…immunisation is about just giving children injection against diseases”…f2RS2.

### Masculinity (attitudes of fathers/husbands)

Attitudes of fathers/husbands/men can help or hinder uptake of childhood vaccines. Household decision-making in the study setting was found to be significantly patriarchal, including on healthcare issues such as childhood immunisation. Thus, immunisation of children depends on the consent or permission of the father/husband, which is significantly linked to his attitude towards immunisation. More importantly, the socio-cultural belief system is crucial to this attitude.

> “You must tell your husband what is happening to the baby”…f1RS3. “We must be carried along”…f2RS4. “…he is the one who takes the major responsibility for the children…his decision is the most important”…f2RS4. “As for me, if anything happens without his consent, it means that I will be held responsible”…f2RS5; “I have to ask permission from him as the head of the family” …f3RS6.

### Gender disparity

In generally, due to several influences, including the culture of gender roles, male children tend to receive a higher level of protection, including life preservation through immunisation, compared to female children. In the same vein, the study found a perception of gender superiority, driven by the knowledge-gap that male children have stronger body immune systems and therefore require little or no immunisation. Therefore, the motivation for immunisation of female children compared to male children is predominantly driven by the notion that females are the weaker gender and therefore require some protective measure than the stronger males.

> “Within the African culture, we put boys first…let’s protect the men more because it is my boy child”…f4RS7; “In the Igbo cultural setting, after you have given birth to six females, he will not even give you the attention to take them for immunisation. But immediately you give birth to a male child, that day they will ask you if you won’t take that child for immunisation. And they will focus on the boy child to protect him from diabolic, physical or other harm”…f3RS6; “Boys are stronger than girls in terms of contracting diseases…they send more of their girls for immunisation than the boys, because their bodies are susceptible to diseases, while boys are stronger because of their immune system”…f2RS7.

### High level of misinformation about vaccines and immunisation

Misinformation associated with vaccines is high among caregivers, especially conspiracy theories, such as the perception of harmful chemicals in vaccines and are deliberately designed to reduce Africa’s population. Others perceived immunisation as a by-pass for family planning therapy. This has created uncertainties, loss of confidence and concerns about the safety of vaccine in general. These behaviour drives vaccine hesitancy and low childhood immunisation uptake in the region.

> “I had a relative…it paralyzes one of her hands. So, I was actually scared and my husband was scared too”…f4RS6. “I am keen on the issue of the vaccines they are bringing to Africa…people are discouraged because of fear of female child infertility as a result of the vaccines”…f1RS3; “People say it has chemicals that are harmful to children…somebody say the immunisation in America is different from the immunisation in Nigeria”…f4RS3”.

### Poor immunisation behaviour despite positive health-seeking attitude

Using caregivers’ intention to vaccinate against Hepatitis B as a measure or indicator of their own vaccination behaviour, there was a significantly higher interest in vaccination against VPDs, including hepatitis B. However, despite this positive attitude or intention, there is low level of hepatitis B vaccination uptake (a measure of vaccination behaviour) among caregivers; driven largely by inadequate information or knowledge.

> “Yes, I would like to receive a Hepatitis B vaccine…but I have not had the opportunity to receive it…f2RS2. “I am very open to it. They brought it to my church, but the crowd was just too much” …f3RS5. “It is very good to take the Hepatitis B vaccine, but in this community, the awareness is very poor. If you ask them what Hepatitis B is, they do not know” …f3RS6.

### Negative impact of COVID-19 pandemic

The COVID-19 pandemic further exacerbated concerns about the safety of vaccines in general, including routine childhood immunisation. Measures introduced to prevent community transmission, such as lockdowns and the perceived corrupt leadership in handling the pandemic, the misinformation about COVID-19 itself, fear of contracting the virus during hospital visits, and the heightened debates about the COVID-19 vaccine affected other healthcare services, including the childhood immunisation service.

> “People believe that the vaccines coming into Nigeria have been mishandled…they are bringing the fake ones into Nigeria”…f1RS3. “COVID-19 has really affected my attitude towards taking any vaccine. From that time till now, when I go to the hospital, they tell me to take any vaccine, no”…f2RS3. “The COVID-19 vaccine caused so many problems, because of the misconception about COVID-19, it made some homes not to take their children for immunisation anymore”…f3RS6.

### Other factors driving low childhood immunisation demand

(1) Cost of immunisation: Exploitation of caregivers

Routine childhood immunisation is free, but the study found that caregivers are exploited by healthcare workers for services in most healthcare facilities.

> “The hospitals charge us for immunisation (spending over ₦30,000 per month), while the government says immunisation is free…so parents who cannot afford it will not immunize the child”…f1RS1. “The money that they asked me to pay was one of the drawbacks, even though I am willing to vaccinate my child”…f1RS 6. “In my experience, you pay for your own vaccines, perhaps this action is not from the government, but the healthcare workers just wanted to extort money for themselves.”…f3RS7.

(2) Non-availability or stock-outs of vaccines

Non-availability or stock-outs of vaccines, especially on immunisation days in some healthcare facilities, contributed significantly to incomplete immunisation and to the inherently complacent immunisation behaviour associated with vaccine hesitancy.

> “Sometimes when you go to the clinic, they don’t have vaccines”…f1RS1. “There are so many health centres that do not offer immunisation”…f1RS4. “My child collects some, but some they don’t have.” …f3RS1.

(3) Ineffective communication about immunisation schedule and reminder system

The inability to effectively communicate information about immunisation, especially the schedule, results in a lack of synchronisation between immunisation providers and the communities/caregivers, which has a negative impact on caregivers’ engagement. Some caregivers also fail to immunize their children because they forget and there is no reminder strategy to help them. This is prevalent during festival and among frequent travellers or nomadic families.

> “For me, I don’t open my doors to the so-called mobile vaccinators, because we are not sure if they are really from the Ministry of Health, because, they would have announced it on the radio”…fR3S4. “The way someone communicates something to you determines how far you’ll go with that information.” …f4RS2; “I forgot because there was no one to remind me…it was December and we were preparing to travel home”…f2RS7.

(4) Religious influence

Religious beliefs were a counterproductive factor in vaccine uptake.

> “One of the reasons is religion, especially denominations within Christianity” …f3RS3. “I didn’t see Jesus Christ vaccinated and I didn’t see immunisation in the Bible” …f4RS7.

(5) Poor attitude of healthcare workers

A significant number of healthcare workers exhibit unprofessional behaviour towards caregivers. More often than not, healthcare workers have unempathetic, rude and exploitative tendencies, which discourages caregivers from visiting immunisation centres.

> “Another thing that discourages parents from going to vaccinated their children is the attitude of the healthcare workers. Some of them are just so bad to us” …f3RS6.

(6) Lack of relationship and trust between healthcare workers and the communities

Where the relationship between healthcare workers and local communities is not characterized by trust, the uptake of healthcare services including immunisation, is negatively affected.

> “The lack of relationship between the healthcare workers and the communities…trust is key”…f3RS5; “We just go to the public and pick anybody who is handy to do the job without training them”…f4RS1. “Who and where the healthcare worker come from matters” …f4RS6.

(7) Fear of Adverse Events from Immunisation (AEFI)

The experience of adverse events from immunisation is a significant factor in immunisation uptake, especially when combined with a lack of prior awareness on the subject. Health scares caused by AEFI make caregivers reluctant to further expose their children to immunisation.

> “Another thing is the fear of AEFI, it makes them not to take it”…f3RS2; “…because after they give the children the vaccine, some children, after a few days, they will become thin, they will cry all night, some for days” …f4RS3.

(8) Insecurity

There is a relationship between general insecurity and low immunisation coverage due to experience or fear of harm. This has hampered immunisation coverage, particularly in hard-to-reach communities served by mobile clinics and vaccinators. It prevents households from opening their doors to mobile vaccinators who go door-to-door to immunise this group and nomadic communities.

> “Just like my brother said about insecurity, these days, not everyone who knocks at your gates is who you want to open the gates for”…f2RS5; “I think the challenge is insecurity in most areas, both urban and rural…you see children being raped, as a result of that people feel that the hospital is not safe either”…fRS6.

## DISCUSSION

The primary aim of this study was to assess the factors influencing caregivers’ immunisation decision-making for children under-5, and to understand the reasons that shape these behaviours. As visualised in Figure 1, the study findings show that caregivers’ immunisation decision-making are mainly motivated by: inadequate knowledge about childhood immunisation, especially about vaccine-preventable and non-vaccine-preventable diseases; masculinity (attitude of fathers/males can help or hurt childhood immunisation); the child’s gender (gender disparity – weaker versus stronger sex); high levels of misinformation or disinformation about immunisation, especially the perception of harmful chemicals in vaccines designed to sterile the population (i.e., family planning through the back door); poor immunisation uptake despite positive health-seeking attitudes; and the spillover effects of the COVID-19 pandemic, which further exacerbate concerns about vaccine safety. Other factors include exploitation of caregivers by healthcare workers; non-availability or frequent stock-outs of vaccines, leading to complacent behaviour associated with vaccine hesitancy; ineffective communication of the immunisation schedule and poor reminder systems; religious beliefs; poor attitudes of healthcare workers; lack of trust between healthcare workers and the host communities; fear of AEFI and general growing insecurity.

**Figure 1:**
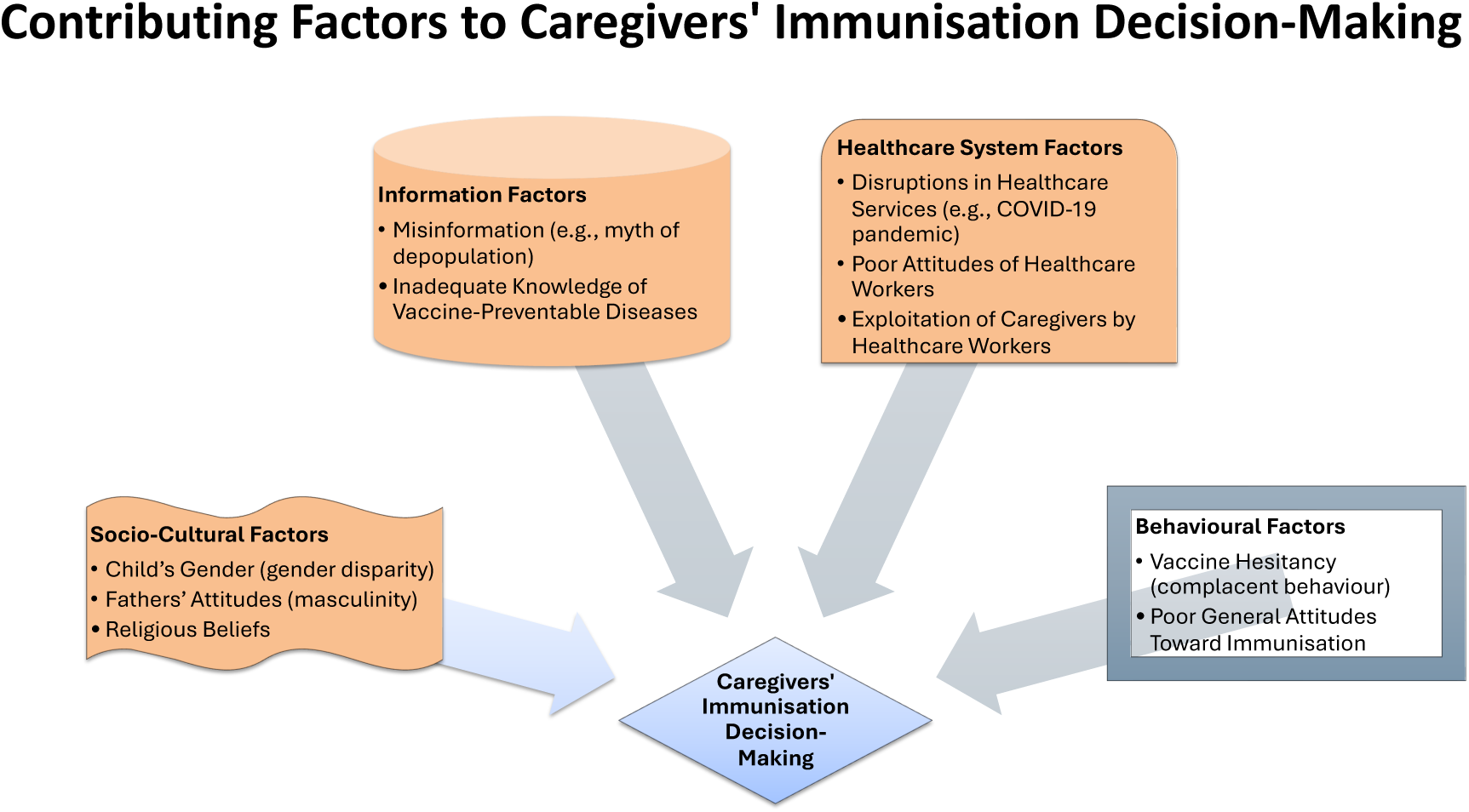
Contributing factors to caregivers’ immunisation decision-making

### Closing the immunisation knowledge-gap

Childhood immunisation appears to be a priority for caregivers, as the study found an increased sense of protection by caregivers for their children’s health, due to its prophylactic capacity, hence the importance attached to immunisation despite the cost (financial burden), AEFI, and other barriers. However, inadequate knowledge about immunisation continues to hamper uptake in low-resource settings, especially in Nigeria. Previous studies have shown that this affects the ability of caregivers to distinguish between vaccine-preventable and non-vaccine-preventable diseases [14,21]. Unfortunately, this study shows that this phenomenon continues to affect immunisation coverage in the country, post COVID-19 pandemic. This knowledge-deficiency is a breeding ground for lack of confidence, trust and consequently, complacent behaviour among caregivers.

Caregivers’ confidence in vaccines is likely to be diminished if, despite immunisation, children continue to suffer or die from diseases from which they thought they were protected, because the knowledge available to them suggests that once a child has been vaccinated, he or she is protected from diseases or death. This misconception is a recipe or breeding ground for vaccine hesitancy. Also, it is more likely to lead to complacent behaviour, based on the perception that immunisation is just a lie, ineffective or after all unimportant because it does not prevent all childhood diseases, as caregivers once believed. “In my own environment I see that some people still don’t accept it. They don’t consider it important”…f3RS5. The per-ception among caregivers that immunisation is a blanket insurance or licence against disease (vaccine-preventable or not) or death is misleading and needs to be corrected urgently.

Therefore, it is very important to provide the population as a whole with adequate knowledge about vaccines and immunisation is very important. Community health education interventions are needed to educate caregivers and the population in general about diseases that are preventable by vaccines and those that are not. Similarly, retraining of healthcare workers, who are the frontline health educators at the community healthcare facility level, should be paramount to convey the accurate efficacy of vaccines. Individuals and groups who thrive on misinformation, disinformation or conspiracy theories about immunisation can easily exploit such knowledge-gaps to sow seeds of mistrust about immunisation.

Given that the study found that a significant proportion of the population still lack critical information about immunisation, especially in rural areas, issues related to inadequate information or low levels of awareness, leading to misconceptions about the purpose of vaccines, affect caregivers’ engagement and, inevitably, vaccine uptake. “A lot of people are not aware…in rural areas; they don’t even know they are supposed to take these vaccines” …f1RS3. For immunisation campaigns to be effective, multiple media of information dissemination sources should be used, especially visual communication [20]. Campaigns should be detailed with specific frameworks about what vaccines are, what they do, what they cannot do, and where to get them. “Create more awareness just to tell people about it and the right place to get all these things…I think that will help lots of people to develop the confidence to begin to take them”…f2RS2.

### Impact of vaccine misinformation or rumours

Misinformation appears to have a dominant effect on caregivers’ immunisation behaviour. Misinformation about vaccines being harmful or containing harmful chemicals, and inaccurate information and communication about AEFI in children, is high among caregivers and negatively affects their attitudes towards immunisation, especially in rural areas. “I was told that if I get vaccinated, if I get pain in my arm, I might have to have it amputated”…f1RS3. “So many people you see their leg. They will say it is the result of the immunisation they took” …f4RS4. Misinformation attributed to AEFI is associated with unexplained or poor communication of the expected or likely adverse events that predispose the circumstances, conditions and events surrounding childhood immunisation. AEFI should be a critical component of antenatal services to ensure that caregivers are adequate educated about AEFI. This activity must also be extended to Traditional Birth Attendants who do not interface with the healthcare facilities, to avoid a gap in the community healthcare continuum. When information about adverse events occurs, prior knowledge of the symptoms of AEFI and possible self-agency in responding to them helps to avoid the gaps filled by misinformation about AEFI, such as those expressed by the participants above.

Also, there was a high level of misinformation associated with the vaccines themselves among caregivers (both fathers and mothers), especially conspiracy theories such as the presence of harmful chemicals in vaccines that are intended to be used for population control or that may compromise future reproductive health of children. Vaccine-related misinformation creates uncertainties and fears about the safety of vaccines in general, which in turn affects behaviour toward immunisation. The study shows that one of the clear drivers of vaccine misinformation is the lack of effective communication about vaccines, which leaves room for conspiracies and rumours to thrive.

In general, rumours thrive where the appropriate medium or media to disseminate the right information is dysfunctional or not engaged. Before people are inundate with fake news about vaccines, it is imperative to reach them with the right information that will promote positive vaccine behaviours, especially when caregivers are still pregnant (i.e., during the antenatal period). The lack of effective strategies for communicating information about vaccines and immunisation to caregivers, especially using health informatics, has remained a sore point in the functioning of the health systems of most developing countries including Nigeria.

One of the insinuations driving the de-population perception or misinformation is the low uptake of family planning services and the aggressive nature of the promotion of this healthcare intervention in the SSA region and Nigeria in particular. There is a perception that family planning as a public health intervention is un-African and was designed to target population growth in the region [37]. This perception is widely held by religious and traditional leaders [14,30]. Thus, immunisation as a public health service becomes an unintended victim of this push-pull conundrum in the family planning debate. People perceived immunisation as an indirect way of sterilising children’s reproductive systems, thereby reducing their ability to have children in the future [14,21,22,30].

### Debunking misinformation

The mythical theory of de-population has caused significant setbacks to various healthcare programmes in Africa, especially immunisation [3,38]. Similarly, several attempts and inter-ventions have been made to debunk and re-educate the population without significant success, as equally observed in this study [39,40]. However, it is noteworthy that these debunking methods may not have been grounded on any evidence-based theories, hence the ineffectiveness of current interventions to address misinformation.

This study found the “truth sandwich” theory, developed by the Robert Koch Institute (RKI) for debunking common misinformation about immunisation, to be a critical asset or tool [41,42]. Although the truth sandwich model was designed for patient consultation scenarios, however, it is well suited to interventions targeting misinformation in low-resource settings, where trust in healthcare workers (a reliable source of immunisation information) has been shown to be very high [15,16,21,22].

First, the *fact or truth* about immunisation must be stated, including that the protection offered by vaccines is safe and effective, although it is not 100% guaranteed and is limited to diseases that can be prevented by immunisation. Interventions must simplify the facts about immunisation to fit with the established knowledge about the effectiveness of vaccines or a particular vaccine that is not controversial, if such a scenario exists. Secondly, the *myth, conspiracy or misinformation* about immunisation must be identified and echoed. In this case, as identified in this study, misinformation that childhood vaccines contain family planning medications or therapies and are designed to impair the reproductive capacity of the African population needs to be mentioned as a familiar narrative. Third, the *fallacies* should be explained, in particular how and why the misinformation is not only inaccurate but misleading, and its origin and/or possible motives, if known to be factual. Finally, the interventions targeting misinformation must repeatedly reinforce the *facts* until they provide an alternative causal explanation and remain the only information taken away.

The truth sandwich model should go a little further by *explaining the damage done* by the myth or the consequences of the myth (e.g., low vaccine uptake makes populations vulnerable to disease outbreaks). I.e., use practical examples of the real-world consequences of the damage caused by vaccine myths and misinformation. Myths and misinformation are major ingredients that fuel vaccine hesitancy, leaving populations vulnerable to disease outbreaks, preventable deaths and increased strain on the healthcare systems, especially in low-resource settings. As shown in Figure 3, “*explaining the damage done*” should come before the final layer, i.e., the reinforcement of the *fact* about the safety and efficacy of vaccines.

### Gender as a driver of demand for immunisation

Gender is a strong factor in vaccination decision-making or behaviour in this study setting, but for different reasons. There appears to be a dual approach to immunisation, hence the disparity in coverage observed between boys and girls in this study setting and across SSA. This is dangerous for vaccine uptake and also a recipe for vaccine hesitancy and outbreaks of VPDs.

Although both male and female children have an equal need for immunisation, there is a disparity in immunisation uptake between boys and girls, accentuated by cultural beliefs and perhaps misinformation. It was not surprising to find the child’s gender as a driver of immunisation uptake in Nigeria, given recent evidence suggesting this [14,21]. However, child’s gender as a driver of immunisation uptake in both directions is novel. That is, being a boy as a driver, and at the same time being a girl as a driver of immunisation behaviour. On the one hand, it is not extremely surprising in the study setting (partly due to the influence of culture) that male children receive more protection, including immunisation, than female children. However, it is surprising to find a perception of gender superiority of the male children, as males are perceived to have a stronger immune system and therefore require less or no immunisation. On the other hand, the motivation to immunise female children compared to male children is driven by the notion that females are the weaker sex and therefore need some protection than the supposedly stronger males.

The centrepiece on immunisation, which is dominated by a preference for boys over girls, was found to be primarily because of the guardrail on the economic and financial provider of the households in the community. He needs to be strong and healthy; therefore, if immunisation guarantees this, then he is the priority. The idea of immunisation as a prophylactic or a form of prevention of harm to the head of HHs meant that the egghead of the family had to be at the centre of protection to ensure the economic survival of the HHs.

The gender disparities in immunisation found in this study are significantly driven by cultural and socio-economic considerations, as well as misinformation or inadequate knowledge, which in most cases are not peculiar to immunisation. Therefore, cultural influences and misinformation appear to be the most prominent drivers of gender differences in childhood immunisation uptake. This study is a reminder that the SSA region is still strongly marked by gender inequalities in key areas, which have historically and disproportionately affected women, whether from a social, economic or political perspective. Nevertheless, efforts to close this gap have come a long way, although it is much narrower than it used to be. This study is a reminder that much more needs to be done to close this gap, especially in the area of the healthcare system. Over the past few decades, societies, policies and populations have evolved to overcome gender discrimination against women, but it was known that this also affected immunisation, especially for infants and young girls under-5.

### Masculinity

Fathers can make or mar immunisation uptake. Their individual perceptions and attitudes towards immunisation have a significant impact on whether a child or children within the HHs are allowed to be immunised. More importantly, other factors also influence such attitudes such as the costs involved (men are perceived responsible), mother’s fear of being blamed in the event of an adverse event or incidence (s) in the child that may be directly or indirectly related to immunisation, and the importance of respecting men’s position as heads of the HHs. Immunisation without the father’s permission attract consequences that are socially undesirable. While mothers are more likely to carry out the actual logistics of childhood immunisation, the decision to vaccinate or not to vaccinate depends significantly on how the fathers feel or think about vaccines or immunisation. Therefore, their attitudes play a key role in the decision and influence whether the children start, continue or complete the full doses of the childhood immunisation. One caregiver said: “He would be the one to provide the transport or any other fees that I am going to use. So, I will take permission from him because of the money I would collect and other things”…f3RS1.

For socially desirable reasons, a few men shied away from the topic during this study. Nevertheless, the majority of the male participants agreed with the notion that they understand their HHs best and therefore the decision about the HHs rests on their shoulders. There are other male participants who, for socially desirable or being politically correct reasons, do not want to explicitly agree with the deposition of the former: thus, on the one hand, they gave answers suggested that immunisation decisions are mutual between mothers and fathers, and that permission is unnecessary since no harm is intended by immunising the child. However, their explanations were accompanied by a caveat that qualified these socially desirable responses, thus further confirming the assertions of the former. For example, one of them alluded to the following: “It is not necessary, however, anywhere my wife goes, I would have to have my consent”…f2RS6.

The study shows that when fathers’ knowledge and confidence in childhood immunisation is high, mother’s attitudes towards immunisation are generally positive, regardless of their own disposition, but not vice versa. In other words, when fathers’ attitudes towards immunisation are negative, this affects mothers’ ability to immunise, regardless of their own attitudes. So, the fathers provide the necessary support to the mothers, including showing interest in the women’s level of awareness about childhood immunisation and getting personally involved in the whole antenatal and postnatal care activities. Therefore, it is quite evident that lack of knowledge about the importance of vaccines and negative perceptions among the fathers have serious implication for mothers when it comes to childhood immunisation. In addition, the cultural environment for routine immunisation is strongly male-dominated (patriarchal), where men are the head of their HHs and women are expected to submit to their authority. The decision-making of the HHs in the study setting demonstrates the complex cultural influence on behaviour, including for immunisation, which is dominated by masculinity. “The man is the head of the household. He owes his wife protection; his wife owes her husband obedience” [43].

Inadequate knowledge and misconception about vaccines and immunisation among fathers could discourage mothers from seeking immunisation services. Also, when fathers’ perceptions about immunisation are negative, the decision to immunise children could become a colleterial damage in the event of disagreements in the HH. “Sometimes husbands and wives disagree, the husband would think that he is the head, and he decides a lot about what the family should do…he might prevent the wife from taking the child for immunisation.”…f2RS3. Masculinity is therefore a clear threat or barrier to immunisation uptake, especially in Nigeria and more broadly in SSA, so interventions that specifically target men or fathers needs to be promoted and scaled up. In this environment, children whose fathers are hesitant to vaccinate are less likely to start, continue or complete the full course of immunisation.

### Connecting caregivers’ positive attitudes to behaviour

In general, the poor immunisation behaviour observed in the study, despite positive health-seeking attitudes, may be attributable to widespread poor health literacy. The study found dearth of health literacy or a high knowledge gap in the setting, which significantly affects attitudes towards healthcare services including immunisation. This was demonstrated by the low uptake of vaccines (e.g., Hepatitis B) among caregivers, despite positive health-seeking attitudes, mainly because knowledge about immunisation is still relatively poor. The immunisation campaigns may need to be redesigned to improve the self-agency of the population in order to translate positive attitudes into actual behaviour through vaccine uptake. “The method of communicating vaccination to parents matters…”…f4RS1.

### The COVID-19 pandemic factor

The negative impact of the COVID-19 pandemic on immunisation services will take a long time to reverse. One of the healthcare services affected most by the pandemic was childhood immunisation [31]. The pandemic set back progress in immunisation coverage by many years and reset norms about attitudes to vaccines. A major lesson from this study is that the country’s healthcare system needs to develop robust operational guidelines and protocol for pandemic preparedness. This would prevent some of the draconian and non-evidence-based interventions measures during the pandemic that further complicated and almost collapsed the healthcare system in Nigeria. It was a double tragedy in the SSA region: because there was the pandemic on the one hand, and the negative impact of the pandemic response on the other, which further crippled the lives and livelihoods of most communities. These scenarios and the backlash from the population created the fertile ground for the spread of misinformation not only about the COVID-19 pandemic, but also about everything directly and indirectly related to it, such as the healthcare services and vaccines in general.

### Exploitation of caregivers

Exploitation of caregivers at the healthcare facilities requires a special attention. Previous studies have found similar behaviour, yet it persists [14,21]. This has been identified as a major barrier that not only affects access to immunisation, but also promotes psychological constraints to immunisation uptake, especially in low-resource settings [14,21]. While caregivers may be willing to immunise children, constraints such as affordability may be the greatest barrier, especially in low-resource settings such as the SSA region, as explicitly further posited by these participants:

> “The money that they asked me to pay was one of the determinants…despite the fact that I am willing to vaccinate my child”…f1RS 6. “Let government make immunisation services to be free”…f2RS1. “If I carry my child for immunisation and I don’t have money I might not get it. Let them help us and make immunisation free”…f2RS4.

Therefore, addressing the issue of illegal fees or charges or exploitation by the healthcare workers could minimise the turn-off of the willing caregivers.

### Training and retraining of healthcare workers

The healthcare workers are a critical factor in the success of immunisation programmes. Therefore, considerable attention should be paid to the training and retraining of personnel involved in immunisation programmes. Retraining and the establishment of best practice guidelines and protocols in healthcare facilities, in addition to the signing of codes of conduct, should address the incessant poor attitudes and the exploitative tendencies of some healthcare workers observed in this study. In addition to being evidence of a fractured healthcare system that lacks leadership and systemic guidelines, immunisation demand cannot thrive where systemic unprofessional behaviour by healthcare workers towards caregivers is prevalent. Healthcare workers are not only exploitative, but also unsympathetic and often rude to caregivers. This is systemic, i.e., not just limited to immunisation services, and therefore requires a drastic overhaul, especially at the rural level.

### Role of incentives

Incentivising healthcare services has been shown to be successful, especially in low-resource settings [44-46]. Such motivation is needed in this setting where several barriers, including poor attitudes towards healthcare, have limited immunisation uptake despite some positive health-seeking intentions among caregivers. Incentivising caregivers will boost participation in immunisation programmes and is likely to bring the healthcare system back to the drawing board. “…support parents with things like mosquito nets”…f2RS2. “…support parents even if it’s a mosquito net, because the nurses have discouraged many mothers”…f4RS2.

### Reminder system

Immunisation campaigns without an effective reminder system will not adequately address the phenomenon of high dropout, incomplete or zero-doses in Nigeria. Therefore, redesigning or leveraging on various new communication tools to enhance caregiver follow-up and reminders is critical. The use of phone calls, text messages and others reminders are critical tools to help caregivers not only start, but complete the routine immunisation. “I think there should be sending of messages to remind parents…” RS2; “They should call parents” …RS3.

### Heightened insecurity and immunisation demand

The relationship between higher levels of insecurity, lack of trust and fear of safety and low vaccine uptake is increasingly driving negative attitudes towards immunisation. Evidence shows that in settings where violence and victimisation prevalence or widespread, child health and immunisation uptake in particular suffer [19,47]. This is because children of victimised caregivers are less likely to be fully immunised and will avoid visiting public healthcare facilities for care [47]. Therefore, addressing the issue of safety and caregivers’ fear or mistrust of the healthcare system/workers is key to transforming positive healthcare-seeking behaviour to actual immunisation behaviour.

This study is not without its limitations and these should be acknowledged. While the qualitative research approach allows for in-depth exploration of caregivers’ experiences and exposes rich contextual details, the data collected may not be insufficient to generalise the experiences of everyone in the setting or to address the heterogeneity of experiences across communities. In addition, by using FGD, the study may have inadvertently introduced a social desirability bias, capable of jeopardising heterogeneity of caregivers’ views. However, despite these limitations, the cross-sectional nature, including the saturation of the qualitative data presented in this study, and the depth of insights are sufficient for the conclusions reported in the study.

## CONCLUSION

This study was able to establish a relationship between the child’s gender (gender disparity), fathers’ attitudes (masculinity), weaponisation of inaccurate information or misinformation, inadequate knowledge of immunisation, generally poor attitudes towards immunisation, disruptions in healthcare services such as those caused by the COVID-19 pandemic, among others, as fundamental factors in how caregivers make decisions about immunisation in low-resource settings such as Nigeria. Because of this link between gender inequality, masculinity and the weaponisation of inaccurate information in relation to immunisation behaviour, the likelihood that boys will have higher immunisation coverage than girls because of the protection of male children, and that girls will have higher immunisation coverage than boys because of the protection of the weaker sex, are both embedded in a culture of masculinity reinforced by patriarchy.

Therefore, when designing interventions to address the root causes of gender inequity, the attitudes of fathers and the social norms and cultural practices that enable them must be addressed first. Societies with high levels of masculinity are more likely to have lower immunisation coverage among boys when beliefs about immunity are gendered, and lower coverage among girls when beliefs about the weaker sex are high. Nevertheless, the overprotection (via immunisation) based on the cultural practices and norms in Nigeria puts the female child at a disadvantage, as she is less likely to be immunised than the male child. There is a need for new ways of communicating immunisation information to caregivers, including immunisation schedules and the reminders. Current strategies do not appear to be effective. The method of communicating immunisation information to caregivers matters and needs to be integrated into the national framework. Furthermore, the theoretical sandwich model for addressing vaccine misinformation (Figure 2) can be used to address myths and misinformation about vaccines.

**Figure 2:**
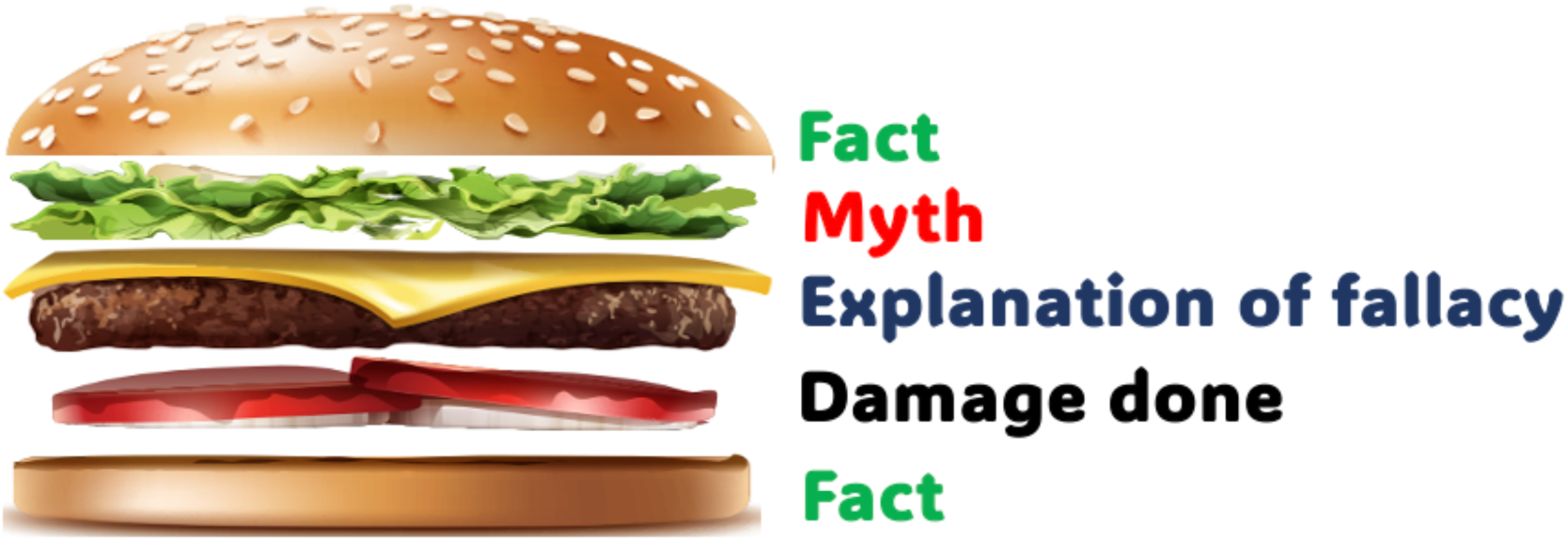
The sandwich model for tackling vaccine misinformation

While the influences of various factors have been reported in this study, further research is recommended, particularly causal (quantitative) research to understand the cause-and-effect or causal relationship between variables such as gender inequality/disparity, misinformation, masculinity, immunisation knowledge, COVID-19 pandemic, among others, and caregivers’ vaccination intention and behaviour or decision-making. This further research would help to explain how changes in a one variable affect vaccination intention and behaviour or decisionmaking, thereby facilitating interventions that could improve immunisation coverage.

## Data Availability

All data produced in the present study are available upon reasonable request to the authors.

## BIBLIOGRAPHY

1. World Health Organization (WHO) (2017). Assessment report of the Global Vaccine Action Plan. Strategic Advisory Group of Experts on Immunization. Retrieved March 1, 2022, from http://www.who.int/immunization/web_2017_sage_gvap_assessment_report_en.pdf.

2. World Health Organization (WHO). (2024b). Over 50 million lives saved in Africa through expanded immunization programme. Retrieved November 21, 2024, from https://www.afro.who.int/news/over-50-million-lives-saved-africa-through-expanded-immunization-programme.

3. Bangura, B. J., Xiao, S., Qui, D., Ouyang, F., & Chen, L. (2020). Barriers to childhood immunization in sub-Saharan Africa: A systematic review. BMC Public Health, 20, 1108. 10.1186/s12889-020-09169-4.

4. Mankhwala, C. S., Chifungo, C., Mzembe, T., Nwira, T., Peterson, M. B., Khundi, M., Madise, N. J., & Chipeta, M. G. (2024). Assessing the resilience of child immunization programmes using geospatial modeling and interrupted time series analysis in Ethiopia and Kenya amidst the COVID-19 pandemic: Tracking coverage and identifying key challenges. BMJ Public Health, 2(1), e00857. https://bmjpublichealth.bmj.com/content/2/1/e000857

5. Adesina, M. A., Olufadewa, I. I., Oladele, R. I., Solagbade, A., & Olaoyo, C. (2023). Determinants of immunization among rural mothers in Nigeria. Population Medicine, 5(September), 22. 10.18332/popmed/171542.

6. World Health Organization & UNICEF. (2019). WHO/UNICEF (WUENIC): Estimates of national immunization coverage. Retrieved January 2, 2022, from https://www.who.int/immunization/monitoring_surveillance/who-immuniz.pdf.

7. World Health Organization (WHO) African Region. (2023). Status of immunization coverage in Africa as of the end of 2022. Brazzaville, WHO African Region. https://www.afro.who.int/sites/default/files/2023-10/Status%20of%20immunization%20coverage_final-compressed_compressed.pdf.

8. Shearer, J. C., Nava, O., Prosser, W., Nawaz, S., Mulongo, S., Mambu, T., Mafuta, E., Munguambe, K., Sidauque, B., Cherima, Y. J., Durosinmi-Eti, O., Okojie, O., Hadejia, I. S., Oyewole, F., Mekonnen, D. A., Kanagat, N., Hooks, C., Fields, R., Richart, V., & Chee, G. (2023). Uncovering the drivers of childhood immunization inequality with caregivers, community members, and health system stakeholders: Results from a human-centred design study in DRC, Mozambique, and Nigeria. Vaccines, 11(3), 689. 10.3390/vaccines11030689.

9. National Primary Healthcare Development Agency (NPHCDA). (2017). National immunization coverage survey (NICS) 2016/2017: National immunization coverage brief. https://www.nigerianstat.gov.ng/nada/index.php/catalog/59/download/574.

10. Lawal, T. V., Atoloye, K. A., Adebowale, A. S., & Fagbemigbe, A. F. (2023). Spatiotemporal analysis of childhood vaccine uptake in Nigeria: A hierarchical Bayesian zero-inflated Poisson approach. BMC Pediatrics, 23, 493. 10.1186/s12887-023-04300-x.

11. Ophori, E. A., Tula, M. Y., Azih, A. V., Okojie, R., & Ikpo, P. E. (2014). Current trends of immunization in Nigeria: Prospects and challenges. PMC Tropical Medicine and Health, 42(2), 67–75. 10.2149/tmh.2013-13.

12. Breakthrough Research. (2020). Routine childhood immunization: Insights for improving malaria, family planning, and maternal and child health outcomes in north-western Nigeria through social and behavior change programming. Programmatic Research Brief. Population Council. https://knowledgecommons.popcouncil.org/departments_sbsr-rh/1543.

13. Betsch, C., Schmid, P., Heinemeier, D., Korn, L., Holtmann, C., & Böhm, R. (2018). Beyond confidence: Development of a measure assessing the 5C psychological antecedents of vaccination. PLoS ONE, 13(12), e0208601. 10.1371/journal.pone.0208601.

14. Adeyanju, Gbadebo Collins, and Cornelia Betsch (2024). Vaccination Decision-Making among Mothers of Children 0 – 12 Months Old in Nigeria: A Qualitative Study. Human Vaccines & Immunotherapeutics, 20 (1). 10.1080/21645515.2024.2355709.

15. Adeyanju, G. C., Engel, E., Koch, L., Ranzinger, T., Shahid, I. B. M., Head, M. G., Eitze, S., & Betsch, C. (2021a). Determinants of influenza vaccine hesitancy among pregnant women in Europe: A systematic review. European Journal of Medical Research, 26(1), Article 116. 10.1186/s40001-021-00553-7.

16. Adeyanju, G. C., Sprengholz, P., Betsch, C., & Essoh, T.-A. (2021b). Caregivers’ willingness to vaccinate their children against childhood diseases and human papillomavirus: A cross-sectional study on vaccine hesitancy in Malawi. Vaccines, 9(11), 1231. 10.3390/vaccines9111231.

17. UNICEF (2022). UNICEF immunization roadmap to 2030. https://www.unicef.org/media/138976/file/UNICEF%20Immunization%20Roadmap%20To%202030.pdf.

18. Galadima, A. N., Zulkefli, N. A. M., Said, S. M., & Ahmad, N. (2021). Factors influencing childhood immunisation uptake in Africa: A systematic review. BMC Public Health, 21(1), 1475. 10.1186/s12889-021-11466-5

19. Adeyanju GC, Schrage P, Jalo RI, Abreu L, Schaub M (2025e) Armed violent conflict and healthcare-seeking behavior for maternal and child health in sub-Saharan Africa: A systematic review. PLoS ONE 20(2): e0317094. 10.1371/journal.pone.0317094

20. Adeyanju GC, Frampton S, Hein C (2025a). Diphtheria and the risk of outbreaks of vaccine-preventable diseases in low-resource settings. Academia Medicine. 2(1). 10.20935/AcadMed7528

21. Adeyanju, G. C., Sprengholz, P., & Betsch, C. (2022a). Understanding drivers of vaccine hesitancy among pregnant women in Nigeria: A longitudinal study. NPJ Vaccines, 7(1), 96. 10.1038/s41541-022-00489-7.

22. Adeyanju, G. C., Betsch, C., Adamu, A. A., Gumbi, K. S., Head, M. G., Aplogan, A., Tall, H., & Essoh, T.-A. (2022b). Examining enablers of vaccine hesitancy toward routine childhood and adolescent vaccination in Malawi. BMC Global Health Research and Policy, 7(1), 28. 10.1186/s41256-022-00251-w.

23. Adamu, A. A., Essoh, T.-A., Adeyanju, G. C., Jalo, R. I., Saleh, Y., Aplogan, A., & Wiysonge, C. S. (2021). Drivers of hesitancy towards recommended childhood vaccines in African settings: A scoping review of literature from Kenya, Malawi and Ethiopia. Expert Review of Vaccines, 20(6), 1–14. 10.1080/14760584.2021.1899819.

24. Wiysonge, C. S. (2019, December 17). Vaccine hesitancy: An escalating danger in Africa. Think Global Health. https://www.thinkglobalhealth.org/article/vaccine-hesitancy-escalating-danger-africa. Retrieved March 2, 2025.

25. Gavi, the Vaccine Alliance. (2020). Gender and immunization. Retrieved February 25, 2022, from https://www.gavi.org/our-alliance/strategy/gender-and-immunisation.

26. Feletto, M., & Sharkey, A. (2019). The influence of gender on immunisation: Using an ecological framework to examine intersecting inequities and pathways to change. BMJ Global Health, 4, e001711. 10.1136/bmjgh-2019-001711.

27. Merten, S., Martin Hilber, A., Biaggi, C., Secula, F., Bosch-Capblanch, X., Namgyal, P., & Hombach, J. (2015). Gender determinants of vaccination status in children: Evidence from a meta-ethnographic systematic review. PLOS ONE, 10(8), e0135222. 10.1371/journal.pone.0135222.

28. Hanifi, S. M. A., Ravn, H., Aaby, P., & Bhuiya, A. (2018). Where girls are less likely to be fully vaccinated than boys: Evidence from a rural area in Bangladesh. Vaccine, 36(23), 3323–3330. 10.1016/j.vaccine.2018.04.059.

29. Cooper, S., Betsch, C., Sambala, E. Z., Mchiza, N., & Wiysonge, C. S. (2018). Vaccine hesitancy: A potential threat to the achievements of vaccination programmes in Africa. Human Vaccines & Immunotherapeutics, 14(10), 2355–2357. 10.1080/21645515.2018.1460987.

30. Adeyanju, G. C., Essoh, T.-A., Sidibe, A. R., Kyesi, F., & Aina, M. (2024). Human papillomavirus vaccination acceleration and introduction in sub-Saharan Africa: A multi-country cohort analysis. Vaccines, 12(5), 489. 10.3390/vaccines12050489

31. Essoh, T.-A., Adeyanju, G. C., Adamu, A. A., Tall, H., Aplogan, A., & Tabu, C. (2023). Exploring the factors contributing to low vaccination uptake for nationally recommended routine childhood and adolescent vaccines in Kenya. BMC Public Health, 23(1), Article 912. 10.1186/s12889-023-15670-6.

32. Sadiq, M., Croucher, S., & Dutta, D. (2023). COVID-19 vaccine hesitancy: A content analysis of Nigerian YouTube videos. Vaccines, 11(6), 1057. 10.3390/vaccines11061057.

33. Hoshaw-Woodard, S. (2001). Description and comparison of the methods of cluster sampling and lot quality assurance sampling to assess immunization coverage. Department of Vaccines and Biologicals, WHO. https://iris.who.int/bitstream/handle/10665/66867/WHO_VB_01.26-eng.pdf?sequence=1.

34. Bennett, S., Woods, T., Liyanage, W. M., & Smith, D. L. (1991). A simplified general method for cluster-sample surveys of health in developing countries. World health statistics quarterly. Rapport trimestriel de statistiques sanitaires mondiales, 44(3), 98–106.

35. Saini, M., & Shlonsky, A. (2012). Methods for aggregating, integrating, and interpreting qualitative research. In Systematic synthesis of qualitative research (Pocket Guides to Social Work Research Methods). Oxford Academic. 10.1093/acprof:oso/9780195387216.003.0002.

36. World Health Organization (WHO) Regional Office for Europe. (2021). Guide to qualitative evidence synthesis: Evidence-informed policymaking using research in the EVIPNET framework. WHO Regional Office for Europe. Licence: CC BY-NC-SA 3.0 IGO.

37. Ouedraogo, L., Habonimana, D., Nkurunziza, T., Chilanga, A., Hayfa, E., Tall, F., Kidula, N., Conombo, G., Muriithi, A., & Onyiah, P. (2021). Towards achieving the family planning targets in the African region: A rapid review of task-sharing policies. Reproductive Health, 18(1), 22. 10.1186/s12978-020-01038-y.

38. Mihigo, R., Okeibunor, J., Anya, B., Mkanda, P., & Zawaira, F. (2017). Challenges of immunization in the African Region. The Pan African medical journal, 27(Suppl 3), 12. 10.11604/pamj.supp.2017.27.3.12127.

39. Oku, A., Oyo-Ita, A., Glenton, C., Fretheim, A., Ames, H., Muloliwa, A., Kaufman, J., Hill, S., Cliff, J., Cartier, Y., Owoaje, E., Bosch-Capblanch, X., Rada, G., & Lewin, S. (2017). Perceptions and experiences of childhood vaccination communication strategies among caregivers and health workers in Nigeria: A qualitative study. PloS one, 12(11), e0186733. 10.1371/journal.pone.0186733.

40. Ekezie, W., Igein, B., Varughese, J., Butt, A., Ukoha-Kalu, B. O., Ikhile, I., & Bosah, G. (2024). Vaccination Communication Strategies and Uptake in Africa: A Systematic Review. Vaccines, 12(12), 1333. 10.3390/vaccines12121333.

41. Lewandowsky, S., Cook, J., Ecker, U. K. H., Albarracín, D., Amazeen, M. A., Kendeou, P., Lombardi, D., Newman, E. J., Pennycook, G., Porter, E., Rand, D. G., Rapp, D. N., Reifler, J., Roozenbeek, J., Schmid, P., Seifert, C. M., Sinatra, G. M., Swire-Thompson, B., van der Linden, S., Vraga, E. K., Wood, T. J., & Zaragoza, M. S. (2020). The debunking handbook 2020. 10.17910/b7.1182.

42. Robert Koch Institute (RKI). (2024). Vaccination myths: Effectively debunking misinformation. https://www.rki.de/EN/Content/infections/immunisation/vaccination_myths/vaccination_myths_content.html. Accessed December 12, 2024.

43. Jousse, L. (2021, May 31). Discrimination and gender inequalities in Africa: What about equality between women and men? Gender in Geopolitics Institute. Retrieved December 11, 2024, from https://igg-geo.org/en/2021/05/31/discrimination-and-gender-inequalities-in-africa-what-about-equality-between-women-and-men/#_ftn2.

44. Kiwanuka, S. N., Babirye, Z., Kabwama, S. N., Tusubira, A. K., Kizito, S., Ndejjo, R., Bosonkie, M., Egbende, L., Bondo, B., Mapatano, M. A., Seck, I., Bassoum, O., Leye, M. M. M., Diallo, I., Fawole, O. I., Bello, S., Salawu, M. M., Bamgboye, E. A., Dairo, M. D., Adebowale, A. S., Afolabi, R. F., & Wanyenze, R. K. (2024). Health workforce incentives and disincentives during the COVID-19 pandemic: Experiences from Democratic Republic of Congo, Nigeria, Senegal, and Uganda. BMC Health Services Research, 24, Article 422. 10.1186/s12913-024-06899-3.

45. Okereke, E., Eluwa, G., Akinola, A., Suleiman, I., Unumeri, G., & Adebajo, S. (2021). Patterns of financial incentives in primary healthcare settings in Nigeria: Implications for the productivity of frontline health workers. BMC Research Notes, 14. 10.1186/s13104-021-05671-z

46. Abduljawad, A., & Al-Assaf, A. F. (2011). Incentives for better performance in health care. Sultan Qaboos University Medical Journal, 11(2), 201–206. 10.18295/squmj.2011.11.02.012.

47. Schaub, M., Adeyanju, GC., Abulfathi, AA., Bello, MM., Kasserra, L., Kwaku, AA., Jalo, MI., Mahmud, A., Schrage, P., Jalo, RI., Abreu, L (2025). Maternal and Child Healthcare-seeking among Victims of Violence in Armed Conflict: A Matched Case-control Study in Northeast Nigeria. BMJ Global Health. doi: 10.1101/2025.03.11.25323760

